# Effects of Electrical Pulse Polarity Shape on Intra Cochlear Neural Responses in Humans: Triphasic Pulses with Anodic and Cathodic Second Phase

**DOI:** 10.1101/2020.12.07.20244012

**Authors:** Katharina Kretzer, David P. Herrmann, Sabrina H. Pieper, Andreas Bahmer

## Abstract

Modern cochlear implants employ charge-balanced biphasic and triphasic pulses. However, the effectiveness of electrical pulse shape and polarity is still a matter of debate. For this purpose, in a previous study (Bahmer & Baumann, 2013) electrophysiological and psychophysical measurement after triphasic pulse stimulation with *cathodic* second phase was determined. Depending on the pulse shape configuration, the stimulation effectiveness differed similarly for electrophysiological and psychophysical measurements. However, the experiments were limited to stimulation pulses with *cathodic* second phase.

In this study, *cathodic and anodic* second phase stimulation was applied. Evoked compound action potentials (ECAPs) and psychophysical responses were recorded in eleven cochlear implant recipients (SYNCHRONY/SONATAti100/PULSARci100 devices, MED-EL Innsbruck). We compared the strength of the ECAP responses with individual psychophysical threshold levels depending on the pulse shape.

Results for pulses with *cathodic* second phase showed the weakest ECAP response and highest psychophysical thresholds for symmetric triphasic pulse shapes, and the strongest ECAP response and lowest psychophysical thresholds for biphasic pulses. The ECAP responses for *anodic* second phase differed from the results of triphasic stimulation with *cathodic* second phase. The U-shape of the ECAP response with increasing phase amplitude ratio (PAR) for *cathodic* second phase could not be observed for the *anodic* second phase. Instead, a flat curve was observed. In contrast, psychophysical threshold curves with increasing PAR were similar between *cathodic* and *anodic* second phase stimulation.

## Introduction

Cochlear implants (CIs) can substitute damaged hair cells in the inner ear of patients with severe to profound sensorineural hearing loss. To ensure patient safety, electrical stimulation have to be charge-balanced. Therefore, most modern CI systems emit pulses consisting of two phases of opposing polarity, i.e. biphasic pulses.

Both polarities of a pulse (anodic and cathodic), can depolarize nerve fibers and generate action potentials (van Wieringen et al., 2008). Results of studies with human CI users showed a more effective stimulation of neurons with anodic pulses compared to cathodic pulses (Bahmer & Baumann, 2013, p. 201; Macherey et al., 2008; Undurraga et al., 2010; van Wieringen et al., 2008). Furthermore, current studies have suggested that this difference in polarity sensitivity may be a correlate of neural health (Carlyon et al., 2018; Hughes et al., 2018; Mesnildrey et al., 2020).

A different kind of charge-balanced pulse is the so-called triphasic pulse, which consists of three phases of alternating polarity. This pulse was originally implemented in CI systems to reduce stimulation artifacts in recordings of neural responses after electrical stimulation (Bahmer et al., 2010; Bahmer & Baumann, 2012a, 2012b). However, triphasic pulses have been shown to be also beneficial in cases of CI users suffering from co-stimulation of the facial nerve, (Bahmer et al., 2017; Bahmer & Baumann, 2016; Braun et al., 2019). To investigate whether these pulses are as effective as biphasic pulses, Bahmer and Baumann (2013) recorded the strength of neural responses to changes in pulse shape and polarities by applying so-called precision-triphasic pulses (in the following abbreviated as p-triphasic pulse). A distinctive feature of this shape is the variably adjustable amplitude of the first and third phase. Since all phases have the same duration, the balance of electrical charges is secured via different amplitudes. Consequently, the sum of the amplitudes of the first and third phase equals the amplitude of the second phase in terms of absolute value. As the fixed amplitude of the second phase has the highest value of all, its polarity can be considered dominant (Carlyon et al., 2013). The adjustable ratio of the amplitudes is defined by the quotient of the amplitudes of the first and second phase, which is referred to as the phase amplitude ratio PAR (Bahmer & Baumann, 2012a, 2012b). Consequently, a pulse with a PAR of 0 or 1 is equal to a biphasic pulse. Bahmer and Baumann (2013) used p-triphasic pulses with a fixed cathodic second phase and altered the PAR from 0 to 1 in steps of 0.1. In this way, they successively changed the pulse shape from a biphasic pulse with a cathodic first phase, through differently shaped triphasic pulses to a biphasic pulse with anodic first phase. ECAPs for each PAR were recorded and response strengths were compared with corresponding psychophysically determined detection thresholds. Their results showed that neural response strength and psychophysical detection thresholds depended strongly on the pulse shape. With increasing PAR starting from 0, the ECAP response amplitudes decreased continuously until a minimum at a PAR of 0.4. With further increase of PAR, the efficiency increased again until the maximum was reached at a PAR of 1. Correspondingly, the psychophysical detection thresholds rose from a PAR of 0 to 0.5 and decreased again to a PAR of 1. Whereas a PAR of 0 caused a considerably lower ECAP response compared to a PAR of 1, no comparably pronounced difference between both PAR could be observed at the level of detection thresholds. The comparison of the electrophysiological and psychophysical measures showed a high correlation.

The previously described study employed only cathodic second phase pulses. This study investigated if these results can be transferred to pulses with anodic second phase. ECAP response strength and psychophysical detection thresholds were determined in eleven CI users after stimulation with p-triphasic pulses with *cathodic and anodic* second phase.

## Material and methods

### Participants

Eleven participants (P1-11) with mean age of 51.5 (standard deviation ±18.5) were included in the data evaluation (see Table 1). Inclusion criteria were a minimum age of 18 years and a duration of at least 12 months between implantation and study participation. Only subjects with implants from the manufacturer MED-EL (Innsbruck, Austria) with auditory response telemetry capabilities (PULSARci100, SONATAti100, SYNCHRONY) were included. Functionally, there are no significant differences between these subtypes of the MED-EL devices. Only for SONATAti100 and SYNCHRONY implants, the stimulation reference electrode is located on the implant housing. The reference of the PULSARci100 implants is an external electrode that is placed under the temporalis muscle of the implantee.

**Table 1.** Demographic data of the participants P1-11 of the study. Abbreviations for implant types: PU = PULSARci100; SO = SONATAti100; SY = SYNCHRONY. Abbreviations for electrode types: Std = Standard; Fsoft = FLEXsoft; F28 = FLEX28. In MED-EL CI systems, the contacts are numbered from apical to basal (1-12). The most comfortable loudness levels (MCL) were tested with a PAR of 0 with the pulse polarity that the participant perceived as louder.

| Participant | Implant type | Electrode type | Electrode contact [number] | MCL (PAR = 0) [CU] |
| --- | --- | --- | --- | --- |
| P1 | SO | Fsoft | 2-6-11 | 633-709-945 (cathodic) |
| P2 | PU | Std | 3-6-11 | 567-756-1115 (cathodic) |
| P3 | SY | F28 | 2-6-11 | 633-1134-1087 (cathodic) |
| P4 | SY | Fsoft | 3-6-11 | 614-756-643 (anodic) |
| P5 | PU | Std | 2-6-11 | 1200-1200-1200 (cathodic) |
| P6 | SY | Fsoft | 2-6-11 | 709-992-1200 (cathodic) |
| P7 | SY | Fsoft | 2-6-11 | 737-1134-1134 (cathodic) |
| P8 | SY | Fsoft | 2-6-11 | 614-709-945 (anodic) |
| P9 | SY | Fsoft | 2-6-11 | 945-1200-851 (anodic) |
| P10 | SY | Std | 2-6-10 | 1200-1200-926 (anodic) |
| P11 | SY | F28 | 2-6-11 | 1200-1200-1200 (anodic) |

### Stimuli

For the stimulation, p-triphasic pulses, which can be selected the in MED-EL CI systems were applied (see Figure 1). The p-triphasic pulses were set to PAR values (definition see introduction) between 0 and 1 with a step size of 0.1. The phase duration and the interphase gap were set to 30 µs and 2.1 µs, respectively, for all tests. The stimulation level was specified in current units (CU), with 1 CU corresponding to approximately 1 µA.

**Figure 1.**
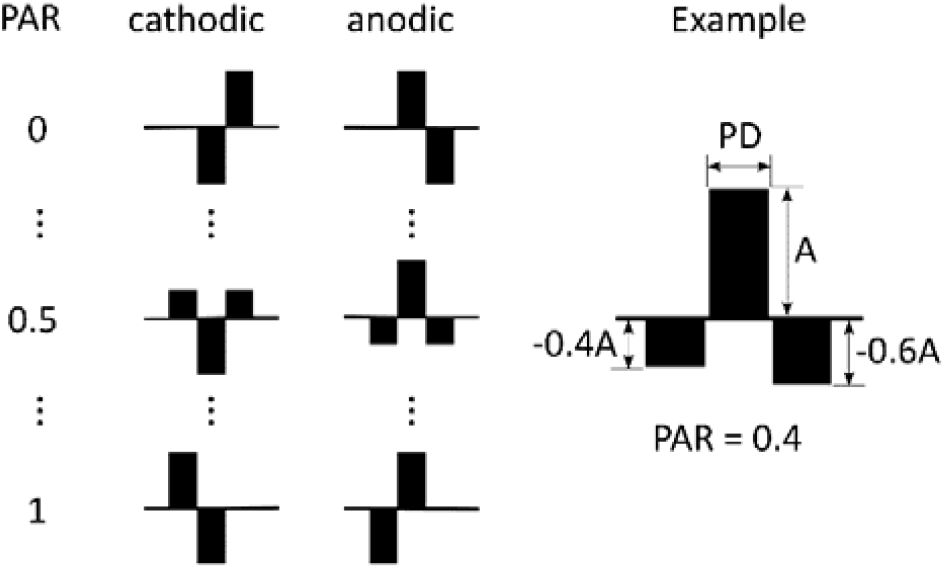
Left: Schematic pulse shapes of precision-triphasic pulses with different polarities and phase amplitude ratios (PAR). Right: example of a precision-triphasic pulse with anodic second phase and a PAR of 0.4. Phase durations (PD) of all phases are identical; the cathodic amplitude of the first phase equals 40% of the amplitude of the anodic second phase (A). To balance the electric charges the cathodic third phase is equal to the difference between the amplitudes of the first and second phase in terms of absolute value.

### Measurement setups and procedures

#### Electrophysiological responses

The setup for the ECAP measurements was similar to the one described by Bahmer and Baumann (2013). A personal computer with 2.5 GHz Dual Core Intel CPU and 8 GB RAM was used. Stimulation and recording parameters were controlled via a graphical user interface programmed in Matlab R2017b (The MathWorks Inc., Natick, USA). A Research Interface Box2 (RIB2; Department of Ion Physics and Applied Physics at the University of Innsbruck, Austria) transferred the stimuli to the participant’s CI via a telemetry coil. ECAPs were recorded and back transferred via the RIB2. A modified forward masking paradigm introduced by Miller et al. (2000) was used for artifact reduction. To improve the signal-to-noise ratio, each sequence of the Miller method was repeated 50 times and finally averaged. The ECAP responses were measured after stimulation from an apical (contact 2 or 3), medial (contact 6), and basal (contact 10 or 11) position on the CI array. Column 6 of Table 1 lists the electrode contact numbers that were used for each participant. To prevent stimulation artifacts, the recording electrode contact was always one position further apical to the stimulating contact.

Prior to the measurements, the stimulation level was determined for each electrode contact of the participant. Therefore, we determined the most comfortable loudness (MCL) at each of the three contacts with a p-triphasic pulse with a PAR of 0 and the polarity that generated the louder percept. The MCL values for each electrode contact and the used polarity for each participant are listed in Table 1. During the actual measurement, the PAR of the p-triphasic pulse was increased from 0 to 1 in steps of 0.1 and the corresponding ECAP responses were recorded. This procedure was conducted using pulses with cathodic and anodic second phase. The data from participants P2, P5, and P7 had to be excluded from further analysis as strong electrical artifacts superimposed their electrophysiological responses. For the participants P1 and P3 only PAR from 0 to 0.9 were tested. The missing values for the PAR of 1 were substituted by the values of the PAR of 0 of the opposite polarity. ECAP response amplitudes and latencies were evaluated for different PARs. The ECAP amplitude was defined as the voltage difference in µV between the negative peak N1 (approximately 0.3 to 0.4 ms after onset of the probe pulse (Brown et al., 1990; Seyle & Brown, 2002)) and the subsequent positive peak P2. The latency was the duration between stimulus onset and N1 peak.

#### Psychophysical detection thresholds

For the determination of the psychophysical detection thresholds, a similar setup was used as described for the ECAP measurements setup. The software “PSYLAB” (Martin Hansen, Institute for Hearing Technology and Audiology, Jade University of Applied Sciences, Oldenburg, Germany) was modified to enable an output with CIs. In contrast to Bahmer and Baumann (2013), in which the experimenter manually determined the detection threshold using an ascending-descending technique, we used a three-interval forced choice adaptive test (Fastl & Zwicker, 2007) to get robust results. For this test, a touch screen presenting three buttons was placed in front of the participant. During one run, three stimulation sequences, each consisting of 50 pulses, were presented in succession. To visually link each sequence to a button, exactly one lit up during the respective presentation. In each run, only one randomly chosen sequence contained an audible stimulus, while the other two sequences contained pulses with an amplitude of zero. The task for the participant was to identify the perceptible sequence and to press the corresponding button after the run was completed. With each correct answer, the stimulation level decreased and with each incorrect answer, the level increased again. The point where a correct answer follows an incorrect one (or the other way around) was defined as a reversal. We used the weighted up-down method after Kaernbach (1991) for the determination of the thresholds. This method enables to determine an individual correct answer probability by introducing different step sizes for upwards (*s*_*up*_) and downwards (*s*_*down*_). We set the step size to 3/1 for *s*_*up*_/*s*_*down*_. This corresponded to a correct answer probability of 75%. The final threshold was determined by averaging the last six reversals.

Detection thresholds for p-triphasic pulses with a cathodic and anodic second phase could be determined for all participants except P6. To keep the test time to a tolerable duration for the participants, we reduced the number of tested electrode contacts to two (apical and medial) and the number of PARs to four (0, 0.2, 0.5, and 0.8).

## Results

### Effects of pulse shape, polarity and contact position on the electrophysiological neural response

Figure 2 shows the latency of all participants (colored lines) in relation to PAR and contact position (colored shapes). The mean value across all participants and electrode contacts (black line), shows that the PAR of 0.2 is related to the longest latency (297 ms) for the cathodic second phase, while a PAR of 0.9 and 1 is related to the shortest latency (272 ms). For the inverse polarity, a PAR of 0.2 showed the longest mean latency (293 ms) and a PAR of 0.8 corresponds to the shortest (254 ms). A Friedman test was used to statistically test the differences between the N1 latency between PARs at the three electrode contact positions and for both polarities. Additionally, Kendall’s W was used as a measure of the effect size. There was no significant difference between the distributions of the N1 latency at the apical and medial contact location for both polarities. At the basal contact, the test found statistically significant differences with a moderate effect size for both polarities (cathodic: p < 0.001, Kendall’s W = 0.456; anodic: p < 0.001, Kendall’s W = 0.383). On the other hand, a post-hoc paired Wilcoxon signed-rank test with adjusted p-values using Bonferroni’s correction for multiple testing showed no significant differences between the N1 latency of the single PARs.

**Figure 2.**
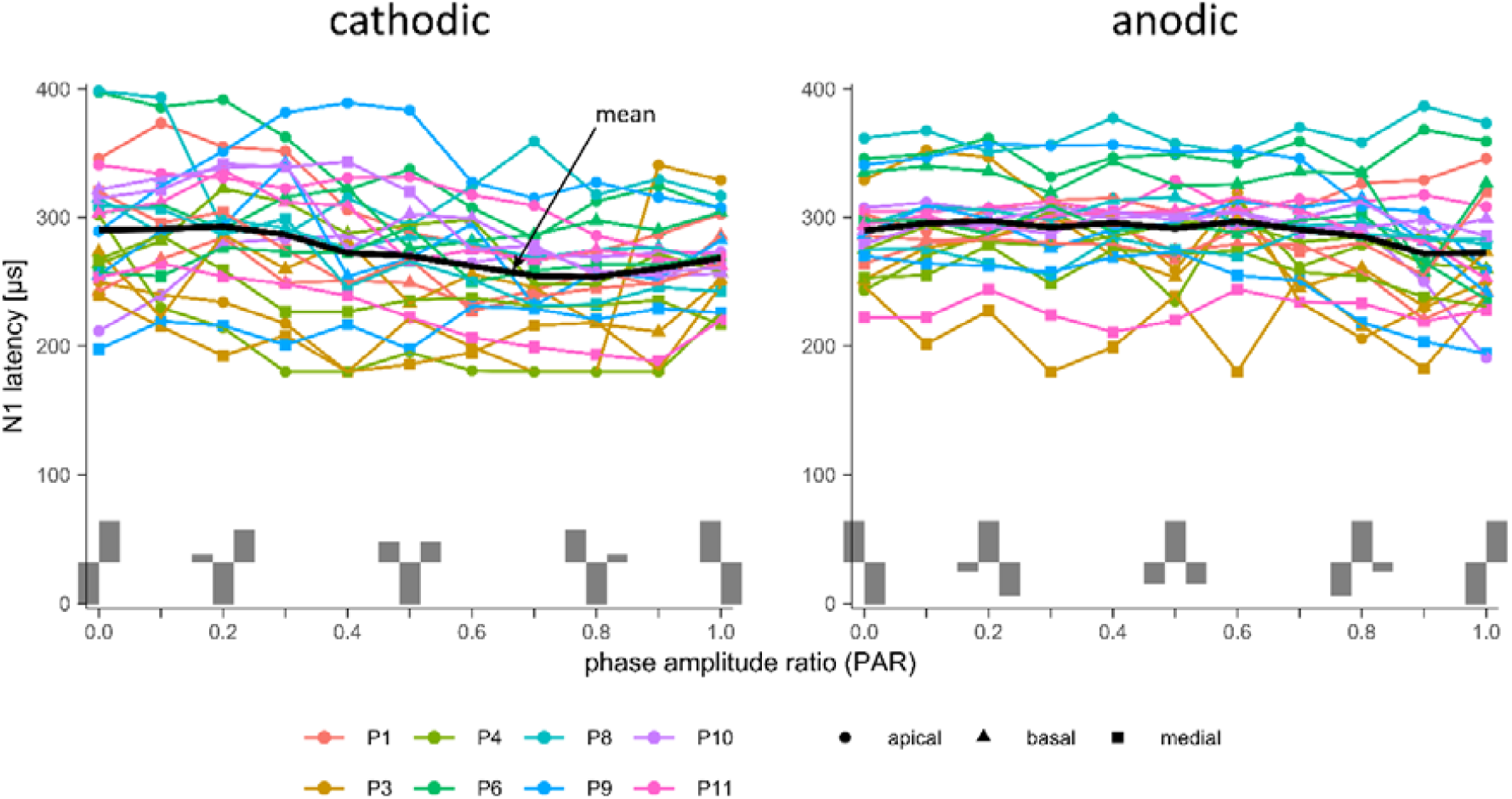
N1 latency of eight participants after stimulation with a p-triphasic pulse with cathodic (left side) and anodic (right side) second phase. The y-axis shows the timespan between the onset of the stimulation pulse and the first negative peak in the recording of the electrophysiological responses, the x-axis displays the phase amplitude ratio (PAR) of the stimulation pulse. Each participant is indicated by a specific color; furthermore, the three tested electrode contact positions (apical, medial, and basal) were given a specific shape. The black line indicates the mean value across all participants and electrode contacts.

For a better visualization of ECAP, amplitudes were normalized. Results of both polarities were pooled then the value 1 was assigned to the highest amplitude and 0 to the lowest amplitude for each participant and electrode contact. The curve of the normalized ECAP amplitudes of all participants (colored lines) and electrode contacts (colored shapes) are plotted for both polarities against the PARs in Figure 3. The black line indicates the median value across all participants and contact positions. On the left side, the median for pulses with cathodic second phase shows a clearly U-shaped curve with a maximum at a PAR of 1 and a minimum at a PAR of 0.4. The results for the anodic second phase on the right side shows a decline of the median normalized amplitude from its maximum at a PAR of 0 to the minimum at a PAR of 0.7. With progression of the PAR until 1 the median value increases again. On average, the amplitudes were highest at a PAR of 1 (biphasic with anodic first phase), followed by a PAR of 0 (biphasic with cathodic first phase). Compared to the results of the cathodic second phase, neural responses to pulses with anodic second phase changed less.

**Figure 3.**
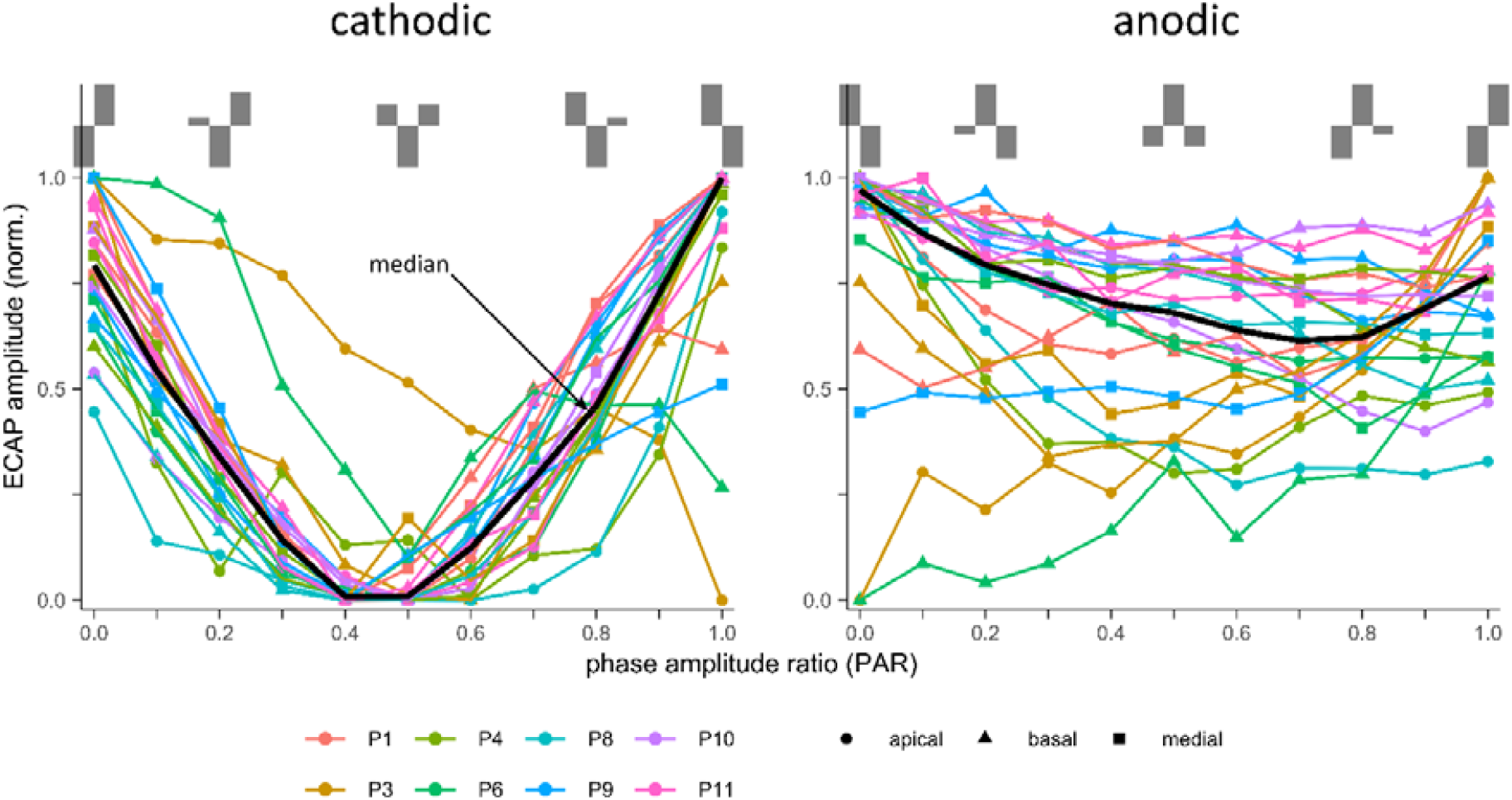
Normalized ECAP amplitudes of eight participants after stimulation with p-triphasic pulses with cathodic (left side) and anodic (right side) second phase as function of the tested PAR. Each participant is presented by a specific color and a specific shape gives each tested electrode contact position. The black line shows the median value across all participants and electrode contacts

Table 2 summarizes the results of the Friedman test testing for differences in the distributions of the normalized ECAP amplitudes between the different PARs at each of the three contact positions and both polarities. Kendall’s W indicates the respective effect size. Except for p-triphasic pulses with anodic second phase at the basal contact position all normalized amplitudes showed statistically significant differences. Kendall’s W indicated a large effect size for the pulses with cathodic second phase in all three electrode contact positions. For pulses with anodic second phase, the effect size was moderate at the apical and medial electrode position and small in the basal position. However, a post-hoc Wilcoxon signed-rank test showed no statistically significant differences between PARs for either polarity.

**Table 2.** Friedman test and Kendall’s W for the normalized ECAP response amplitudes after stimulation with p-triphasic pulses with different polarities and PARs.

| Electrode contact position | Friedman test p-value, Kendall's W |  |
| --- | --- | --- |
|  | cathodic 2<sup>nd</sup> phase | anodic 2<sup>nd</sup> phase |
| apical | < 0.001, 0.674 | < 0.001, 0.398 |
| medial | < 0.001, 0.925 | < 0.001, 0.44 |
| basal | < 0.001, 0.793 | 0.29, 0.149 |

Figure 4 shows the normalized ECAP response amplitudes as median values collapsed across all participants for each electrode contact position. There were statistically significant differences between both polarities for each of the three contact positions (Wilcoxon signed-rank test, p < 0.001). The Spearman’s correlation coefficient showed that the three contact positions were positively correlated within each polarity (see Table 3).

**Table 3.**
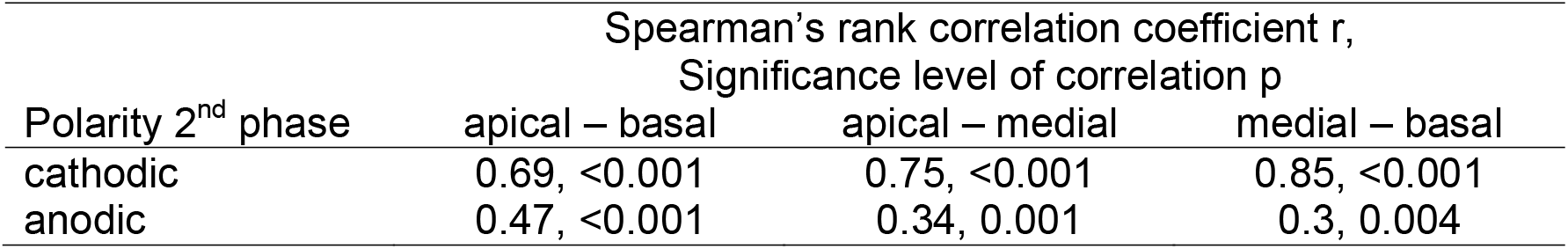
Spearman’s rank correlation coefficients of the normalized ECAP response amplitudes depending on the electrode contact position.

**Figure 4.**
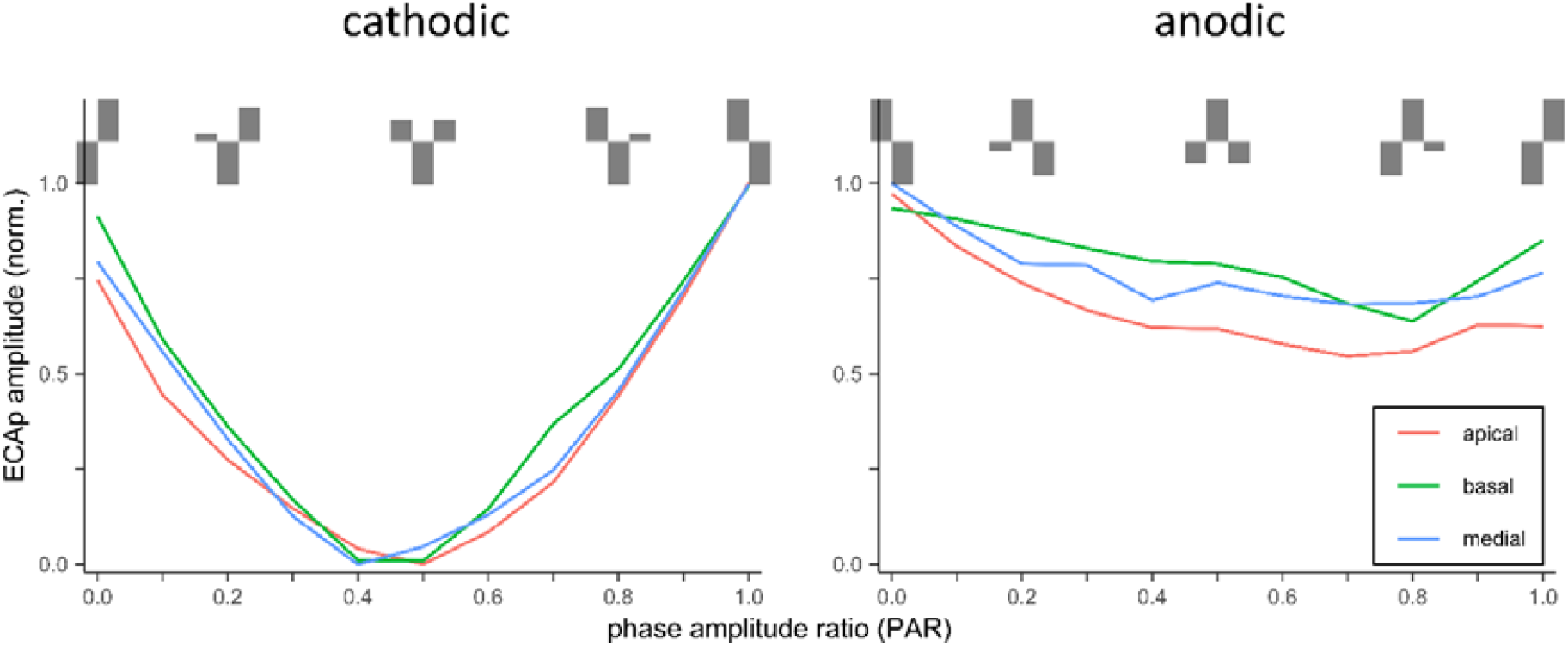
Median values of the normalized ECAP response amplitude collapsed across all participants for each electrode contact position (apical, medial, basal). Left side: p-triphasic pulses with cathodic second phase stimulated responses; right side: responses to pulses with anodic second phase.

### Effects of pulse shape, polarity and contact position on psychophysical detection thresholds

Because of the inter-individual differences in the psychophysical detection thresholds, a normalization was performed similarly as described for the electrophysiological measurements. Analogous to the ECAP measurements, the detection thresholds for a PAR of 1 were substituted by the results for a PAR of 0 of the opposite polarity.

Figure 5 shows the normalized detection thresholds as a function of the tested PARs. For both polarities, the median across all participants and contact positions showed the highest detection values at a PAR of 0.5. The median values decreased successively for PARs smaller or greater than 0.5 until the minima at a PAR of 0 or 1 were reached, respectively. The lower maximum for pulses with anodic second phase at a PAR of 0.5 indicates a more effective stimulation. A Friedman test revealed statistical significant differences with a large effect size between the thresholds of the tested PARs for both polarities and contact positions (see Table 4). A post-hoc Wilcoxon signed-rank test showed significant differences between multiple PARs at the apical contact position and two PARs at the medial position for both polarities (see Table 5).

**Table 4.** Friedman test and Kendall’s W for the normalized psychophysical detection thresholds.

| Electrode contact position | Friedman test p-value, Kendall's W |  |
| --- | --- | --- |
|  | cathodic 2<sup>nd</sup> phase | anodic 2<sup>nd</sup> phase |
| apical | < 0.001, 0.653 | < 0.001, 0.758 |
| medial | < 0.001, 0.593 | < 0.001, 0.685 |

**Table 5.** Detection thresholds. Post-hoc Wilcoxon signed-rank test. p-values were corrected for multiple testing using the Bonferroni’s method.

| Electrode contact position | Wilcoxon signed-rank test, p-values adjusted using<br>Bonferroni's multiple testing correction method |  |
| --- | --- | --- |
|  | cathodic 2<sup>nd</sup> phase | anodic 2<sup>nd</sup> phase |
| apical | PAR 0 vs. 0.2: p = 0.02<br>PAR 0 vs. 0.8: p = 0.02<br>PAR 0.2 vs. 0.5: p = 0.02<br>PAR 0.5 vs. 0.8: p = 0.039 | PAR 0.2 vs. 1: p = 0.02<br>PAR 0.8 vs. 1: p = 0.02 |
| medial | PAR 0 vs. 0.8: p = 0.039 | PAR 0.2 vs. 0.5: p = 0.039 |

**Figure 5.**
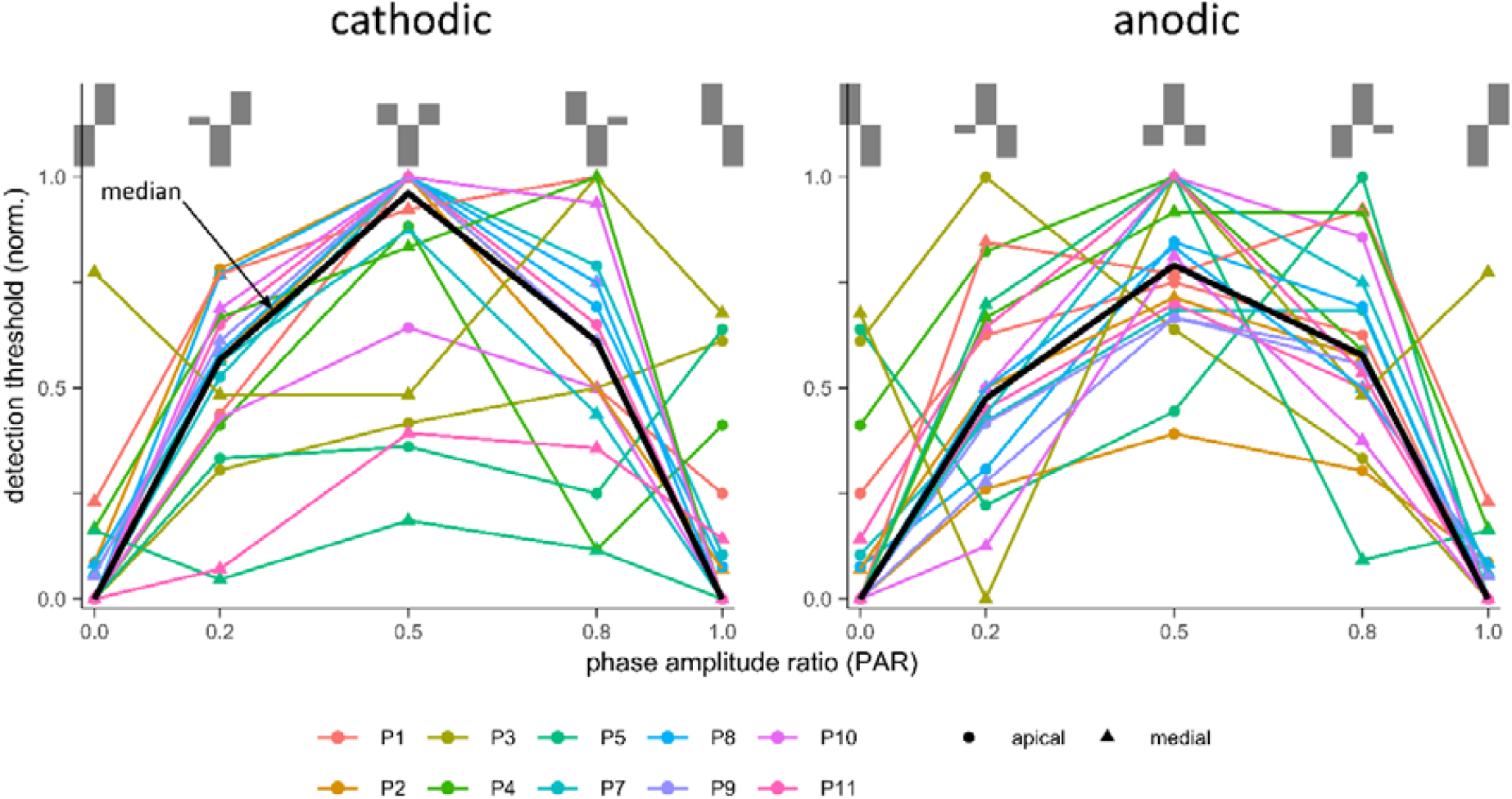
Normalized psychophysical detection thresholds of ten participants stimulated with p-triphasic pulses with cathodic (left side) and anodic (right side) second phase as function of the tested PAR. Each participant is presented by a specific color and a specific shape gives each tested electrode contact position. The black line shows the median value across all participants and electrode contacts.

In Figure 6 the median values of the normalized detection thresholds collapsed across all participants are depicted as functions of the tested PARs for each electrode contact position and polarity (left side: pulses with cathodic second phase; right side: pulses with anodic second phase). The curves are similar both between the contact positions and between the polarities. The median values of both contact positions correlated positively within each polarity (Spearman’s correlation coefficient, r = 0.61 for cathodic second phase, r = 0.54 for anodic second phase, significance level for both correlation coefficients, p < 0.001).

**Figure 6:**
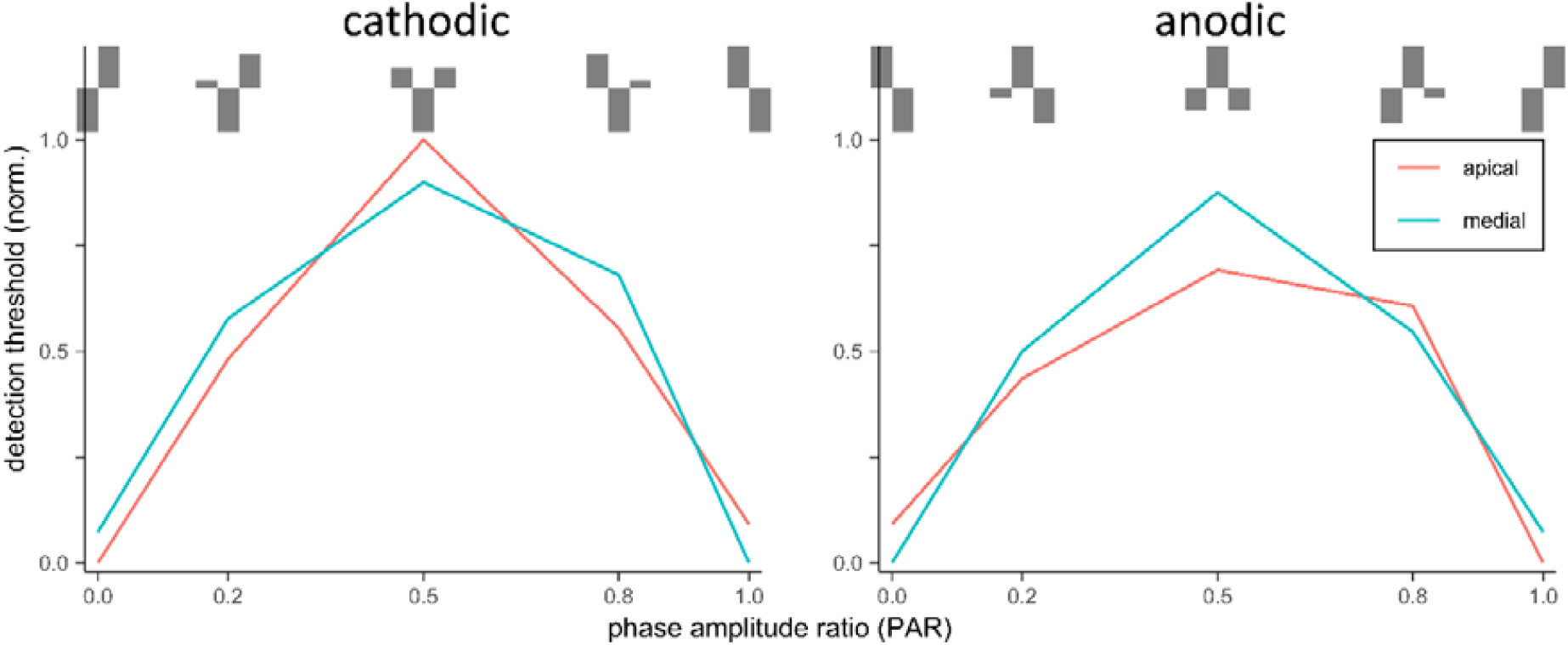
Median values of the normalized psychophysical detection thresholds collapsed across all participants for both electrode contact position (apical, medial). Left side: p-triphasic pulses with cathodic second phase stimulated responses, right side: responses to pulses with anodic second phase.

### Correlation between ECAP responses and psychophysical detection thresholds

To compare the results the electrophysiological measurements and the psychophysical tests, we included only those seven participants who were tested with both methods (P1, P3, P4, P8, P9, P10, and P11). Furthermore, the number of electrode contact positions and the PARs were reduced to the number of electrode contact positions used for the psychophysical tests.

Figure 7 depicts the median values across all participants and both contact positions of the normalized detection thresholds (dashed line) and the inverted normalized ECAP amplitudes (continuous line) in relation to the tested PAR for both polarities of the pulses’ second phase (left: cathodic, right: anodic). For the cathodic polarity, both methods showed a maximum at a PAR of 0.5 and falling slopes with decreasing and increasing PARs. While the inverted ECAP amplitudes reached the minimum at a biphasic pulse shape with anodic first phase (PAR = 1), the detection thresholds were lowest at a biphasic pulse of opposite polarity (PAR = 0). The maxima of the pulses with anodic second phase showed no agreement of ECAP and psychophysical measurement. Similar as for the opposite polarity, the maximum of the normalized detection thresholds reached a maximum at a PAR of 0.5, while the inverted normalized ECAP amplitudes were highest at a PAR of 0.8. Similar as for the opposite pulse polarity, the minima of both methods also differed and showed the same behavior with respect to the biphasic characteristic of the PAR of 0 and 1. A Wilcoxon signed-rank test was used to compare both measures within each polarity. The p-values indicated statistically significant differences within both polarities (p = 0.044 for cathodic second phase, p = 0.002 for anodic second phase). A Spearman’s test showed a positive correlation between both methods within both polarities, which was strong for pulses with cathodic second phase and weak for the opposite polarity (see Figure 7 for Spearman’s r and the corresponding significance level of correlation p).

**Figure 7:**
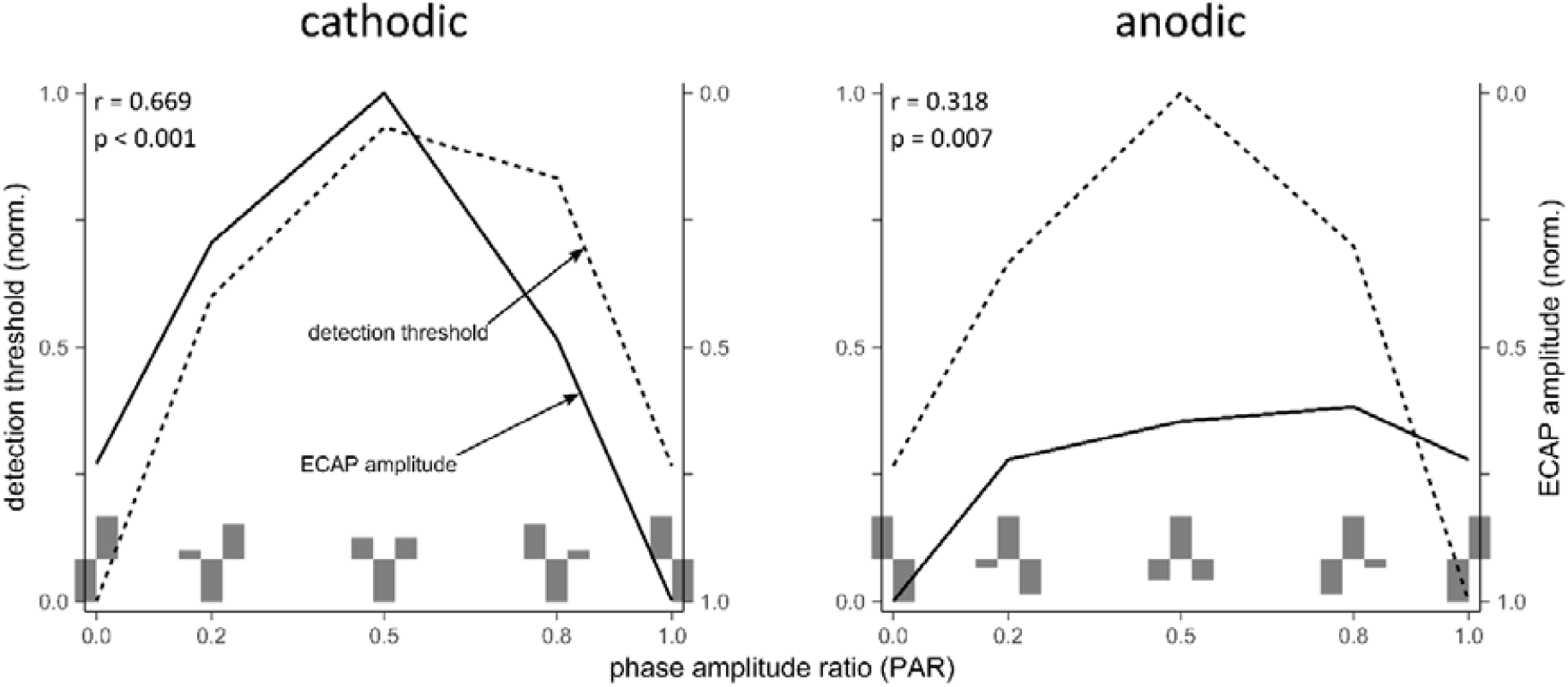
Median values of the inverted normalized ECAP response amplitudes (continuous line) and the normalized psychophysical detection thresholds (dashed line) of seven participants as functions of the tested PAR. Left side: results for p-triphasic pulses with cathodic second phase, right side: results for p-triphasic pulses with cathodic second phase. Spearman’s r and p-values in the upper right corner of each graph indicate the correlation and corresponding significance level of the correlation between both methods within each polarity.

## Discussion

### Effects of pulse shape and polarity

Various studies have described the lower effectiveness of triphasic compared to biphasic pulses (Bahmer & Baumann, 2013; Coste & Pfingst, 1996; Shepherd & Javel, 1999). Our results are in line with these findings. The tested p-triphasic pulses generated smaller ECAP amplitudes and showed higher psychophysical perception thresholds compared to biphasic pulses. The p-triphasic pulses with PARs that resembled biphasic pulses with an anodic first phase evoked the highest ECAP amplitudes and had the lowest threshold levels. Assuming that the first phase of a biphasic pulse and the second phase of p-triphasic pulse represents the dominant polarity in stimulation, the results of this study also reflect a higher sensitivity of nerve fibers to anodic current. This is consistent with the higher sensitivity of human nerve fibers to anodic charge, as described by Macherey et al. (2008).

When the anodic phase varied with the PAR in p-triphasic pulses with a cathodic second phase, the level of the ECAP response varied accordingly. In p-triphasic pulses with an anodic second phase, the ECAP response was more independent of the variation of the cathodic phases since the anodic phase remained constant. Therefore, the anodic phase seemed to have a much stronger influence on the ECAP response. The difference in the neural response properties to anodic and cathodic current is called subsequently *polarity effect*.

### Polarity effect as an indicator for neural health

Current studies investigated the potential relationship between the polarity effect and neural health (Carlyon et al., 2018; Hughes et al., 2018; Jahn & Arenberg, 2019; Macherey et al., 2017). According to these studies, the extent of the polarity effect could depend on the state of neural degeneration and on the simulation level.

#### Neural degeneration

Models of electrical stimulation related the polarity sensitivity of nerve fibers to different sites of action potential initiation. According to these predictions, anodic polarity causes action potentials centrally on the neuron, while cathodic stimulation is more likely to excite its peripheral processes (Joshi et al., 2017; F. Rattay, 1999; F. Rattay et al., 2001; Frank Rattay et al., 2001; Resnick et al., 2018). Demyelination of the peripheral processes or the loss of the latter would therefore result in an increase of the cathodic but not the anodic threshold (Resnick et al., 2018).

Figure 3 depicts the normalized ECAP amplitude of each participant. On average, the p-triphasic pulses with anodic second phase stimulated higher ECAP amplitudes. The extent of the polarity effect showed inter-individual differences. According to the above-mentioned assumptions, a more pronounced difference in ECAP amplitude between p-triphasic pulses with anodic and cathodic second phase would indicate a more advanced degeneration process.

#### Stimulation level

Undurraga et al. interpreted the results of two preceding studies (Macherey et al., 2006; F. Rattay et al., 2001; Frank Rattay et al., 2001) as follows: “At low current levels anodic and cathodic polarities will stimulate the remaining healthy AN [AN = auditory nerve] fibers rather than degenerated ones, which are mainly sensitive to the anodic polarity and present higher thresholds than healthy AN fibers. When the current amplitude increases the cathodic polarity will stimulate the healthy AN fibers and not the degenerated ones, whereas the anodic polarity will stimulate both types of AN fibers, healthy and degenerated.” (Undurraga et al., 2010, p. 159).

The results of Bahmer and Baumann (2013) are in line with this concept. They found an average difference in amplitude of 100 µV for ECAP amplitudes measured at MCL. The p-triphasic pulse with a PAR of 1 (corresponds to a biphasic pulse with anodic first phase) triggered the highest ECAP amplitudes. The determined psychophysical perception threshold showed no difference between polarities. In contrast, Jahn and Arenberg (2019), Macherey et al. (2017), and Carlyon et al. (2018) observed, although not strongly pronounced, both intra- and inter-individually varying polarity effects at the level of perception threshold. As already discussed, the electrophysiologically obtained results of this study showed polarity effects measured at MCL. When analyzed according to Bahmer and Baumann (2013), ECAP amplitude showed an average difference of about 70 µV. On the other hand, the psychophysical perception thresholds revealed no statistically significant polarity effect. However, a visual comparison of the threshold curves between the polarities for individual participants reveals polarity effects of varying intensity to a small degree. Therefore, the results corroborate both concepts to a certain extent.

#### N1 latency

The N1 latencies measured in this study were consistent with observations in previous studies (Abbas et al., 1999; Brown et al., 1990). While Macherey et al. (2008) and Undurraga et al. (2010) reported shorter latencies with anodic stimulation, the comparison in our results showed no statistically significant difference. This may be explained by the differences in the shape of the stimulation pulses, since both studies did not use triphasic stimulation.

#### Comparison between the electrophysiological and psychophysical results

For pulses with cathodic second phase, the inverted ECAP amplitudes and the perception thresholds showed a strong correlation, which is consistent with the results of Bahmer and Baumann (2013). The same comparison for the pulses with anodic second phase showed only a weak correlation (see Figure 7). A possible explanation for the different correlations could be that the polarity effect depends on the stimulation level. According to Undurraga et al. (2010), low stimulation levels mainly stimulate healthy nerve fibers, which react equally strong to both polarities. At higher stimulation levels, degenerated nerve fibers are also excited, which can be stimulated more effectively by anodic polarity (F. Rattay et al., 2001; Undurraga et al., 2010). Since the detection thresholds were determined at low stimulation levels and ECAP responses at high stimulation levels (i.e. MCL), it can be assumed that the differences in the curves for anodic second phase pulses are due to the stimulation levels rather than the test procedures.

## Conclusions

- ECAP responses for anodic second phase were rather constant compared to responses for cathodic seconds phase with increasing PAR
- ECAP response curves for cathodic second phase were u-shaped with increasing PAR.
- Psychophysical detection threshold curves for both anodic and cathodic second phase were inversely U-shaped with increasing PAR.
- The difference between the results of the electrophysiological measurements and psychophysical tests to the anodic and cathodic second phases may be an indicator for the neural health of the stimulated AN fibers.

## Data Availability

All data can be made available by contacting the corresponding author.

## Disclosure

Funding: the Deutsche Forschungsgemeinschaft, Bonn, Germany [BA 4237/3-1], supported this work. There are no conflicts of interest, financial, or otherwise.

## Acknowledgments

We would like to thank the CI recipients for participation. Informed consent was obtained from each participant before participation. The data was acquired under the authorization of the ethics committee of the University of Würzburg (315/15_z).

## Notes

### Competing Interest Statement

The authors have declared no competing interest.

### Author Declarations

The data was acquired under the authorization of the ethics committee of the University of Wuerzburg (file number 315/15_z). Address of the oversight body: Ethik-Kommission der Universitaet Wuerzburg Institut fuer Pharmakologie und Toxikologie Versbacher Str. 9 97078 Wuerzburg, Germany

